# The Sensible Senseless Person: a theoretical framework for clinical assessment of proprioception

**DOI:** 10.1101/2025.10.31.25339219

**Authors:** Yogev Koren, Asaf Cohen, Noy Goldhamer, Lior Shmuelof

## Abstract

Proprioception, the sense of body position and motion, is essential for all motor activities. Position-matching is a common task used to assess proprioceptive acuity. This assessment is limited due to reliance on self-report, and biased performance measures. To support this statement, we introduce the concept of a “sensible senseless person” (SSP), a theoretical construct that lacks sensory perception but is capable of logical inference. This framework allows for critical examination of the validity of conventional performance measures used in position-matching tasks. Our analysis reveals that traditional measures are influenced by the structural parameters of the task, strategic responses, and the prior beliefs of the SSP.

To address these limitations, we propose a novel performance metric: the Guess Index (GI). Unlike existing measures, the GI is designed to be robust against these confounding factors, ensuring that the SSP would consistently receive the same score. To demonstrate the utility of the GI, we applied a simplified, clinically-oriented version of the position-matching task to healthy individuals and persons with stroke. Our findings show that the GI is more sensitive to group differences than traditional measures and is the only metric capable of correctly classifying persons with stroke above chance level.

## Introduction

Proprioception refers to the sense of body position and motion. This sense is essential for all motor activities; without it, movement is poorly controlled, leaving individuals functionally paralyzed [1,2]. Proprioceptors, the sensory organs responsible for proprioception, are distributed throughout the musculoskeletal system [3,4]. Consequently, any damage to the musculoskeletal system, peripheral nerves, or central nervous system (CNS) can result in proprioceptive deficits and impaired control and function. Since poor control and function are not unique to proprioceptive deficits, an accurate diagnosis of proprioceptive deficit is critical for early diagnosis, for monitoring the progression of a disease or condition, and for targeted rehabilitation and the assessment of its efficacy.

Despite the importance of this sense, proprioceptive acuity is difficult to quantify. Most assessments rely on self-reports of body position and movement (i.e., conscious proprioception), which are vulnerable to cognitive interference and bias. Common clinical instruments [5,6] typically use qualitative methods, such as asking a blindfolded participant to identify whether a passively moved joint is flexed or extended, whether a body segment is positioned up or down, or to recall or match a specific joint angle using the contralateral limb. Laboratory-based approaches (see reviews in [7,8]) generally follow similar principles but offer quantitative performance metrics (e.g., [9,10]). However, these methods often require specialized equipment and extended testing durations.

Position-matching is a widely used task for estimating proprioceptive acuity [11]. Employed in both clinical and laboratory settings, the task typically involves a blindfolded participant attempting to match the angle of one joint—such as the right elbow, which is passively positioned by an examiner/robot—with the contralateral joint. The procedure is repeated across a range of joint angles, and the resulting matching errors are used to derive quantitative performance metrics believed to reflect proprioceptive acuity.

Three common measures are typically reported:

1. Accuracy – the mean of all signed matching errors.
2. Mean Absolute Error (MAE) or Root Mean Squared Error (RMS) – the average magnitude of unsigned errors.
3. Precision (also referred to as variable error) – the consistency of matching errors across trials.

A reliable detection of proprioceptive deficits requires measure-specific threshold/s that represent an adequate level of proprioceptive input. One approach to defining such threshold is to estimate the required accuracy in joint position sense based on a known or desired end-effector accuracy in a motor task. For example, consider a biomechanical system consisting of a single 30 cm segment (approximately the length of the lower arm) rotating around a hinge joint (i.e., the elbow). Applying forward kinematics to a joint angle error of 2° translates to an end-effector error of roughly 1 cm. Therefore, if the desired or observed end-effector accuracy is 1 cm, the joint angle estimation error should not exceed 2°. Larger errors would suggest proprioceptive deficit.

An alternative approach, commonly used in medical research, is data-driven. This method involves quantifying matching errors in a reference population, typically healthy young adults, and defining an arbitrary threshold at the upper bound of the 95% confidence interval of their performance.

Each of the aforementioned approaches presents distinct limitations, alongside several shared constraints that will be addressed subsequently. Notably, a central confounding factor, common to both approaches, lies in the objective of the task. In joint angle matching tasks, it is straightforward to compute errors based on discrepancies in joint angles. However, evidence suggests that the CNS may prioritize the elevation angles of limb segments relative to gravity [12], favoring this coordinate frame over that of the anatomical joint angles for internal computations [13–15]. If this is the case, participants may unconsciously interpret the task’s objective as matching limb segment orientation rather than joint angles.

The primary objective of this investigation is to demonstrate that the principal limitation in detecting proprioceptive deficits (using position-matching) stems not from the method employed to establish threshold values, but from the outcome measures themselves. To make this point, we introduce the “sensible senseless person” (SSP), a theoretical construct representing a rational person devoid of sensory input. Within this framework, we reveal the limitations of conventional performance measures in accurately reflecting proprioceptive acuity. Specifically, we demonstrate that the SSP’s performance, on any conventional measure, is influenced by strategic behavior, prior beliefs, and the structural properties of the task itself. Building on this construct, we propose a novel performance metric that accounts for these confounding factors and demonstrate its potential advantages by comparing the performance of healthy adults and persons with stroke (PwS) in a simplified, clinically-oriented version of the position-matching task.

### Theoretical framework

We began this investigation by asking: “What would someone who had lost their sense of position report in a position-matching task?” From a probabilistic standpoint, minimizing matching errors can be achieved by consistently reporting the mean of all tested positions (i.e., joint angles).

Formally, let {*x*_*i*_}_*i*_ represent the set of tested angles and {*y*_*i*_}_*i*_ the corresponding set of reported (matched) angles. In the absence of knowledge (i.e., sensory information) about {*x*_*i*_}_*i*_, the best guess for each {*y*_*i*_}_*i*_ is a constant value *C*. The objective is to minimize the sum of squared errors:

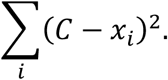

The value of *C* that minimizes this sum is the mean of the tested angles {*x*_*i*_}_*i*_, denoted by 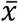. Therefore, the optimal strategy for a person without proprioceptive input would be to report 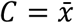 for all trials.

This strategy can be used to predict several characteristics of the distribution of matching errors. Here, the matching error on the *i*th trial, denoted as *e*_*i*_, is the difference between the reported angle, *y*_*i*_, and the tested angle *x*_*i*_ (*e*_*i*_ = *y*_*i*_ − *x*_*i*_). Applying the optimal strategy described above implies that:

1. Errors are expected to be linearly related to the tested angle, following the expression: 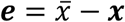
2. Signed errors should be symmetrically distributed around zero (see Figure 1a, red series, for graphical representation), and therefore Accuracy (mean of all signed errors) would equal zero, which could misleadingly suggest perfect performance.
3. The measures Precision and MAE (and RMS) are dependent on the range of angles tested, that is, on the structure of the test itself. For instance, a test using angles of 0° and 90° would yield a much higher MAE (45°) than a test using angles of 30° and 60° (MAE=15°), and both have an accuracy of zero.

**Figure 1.**
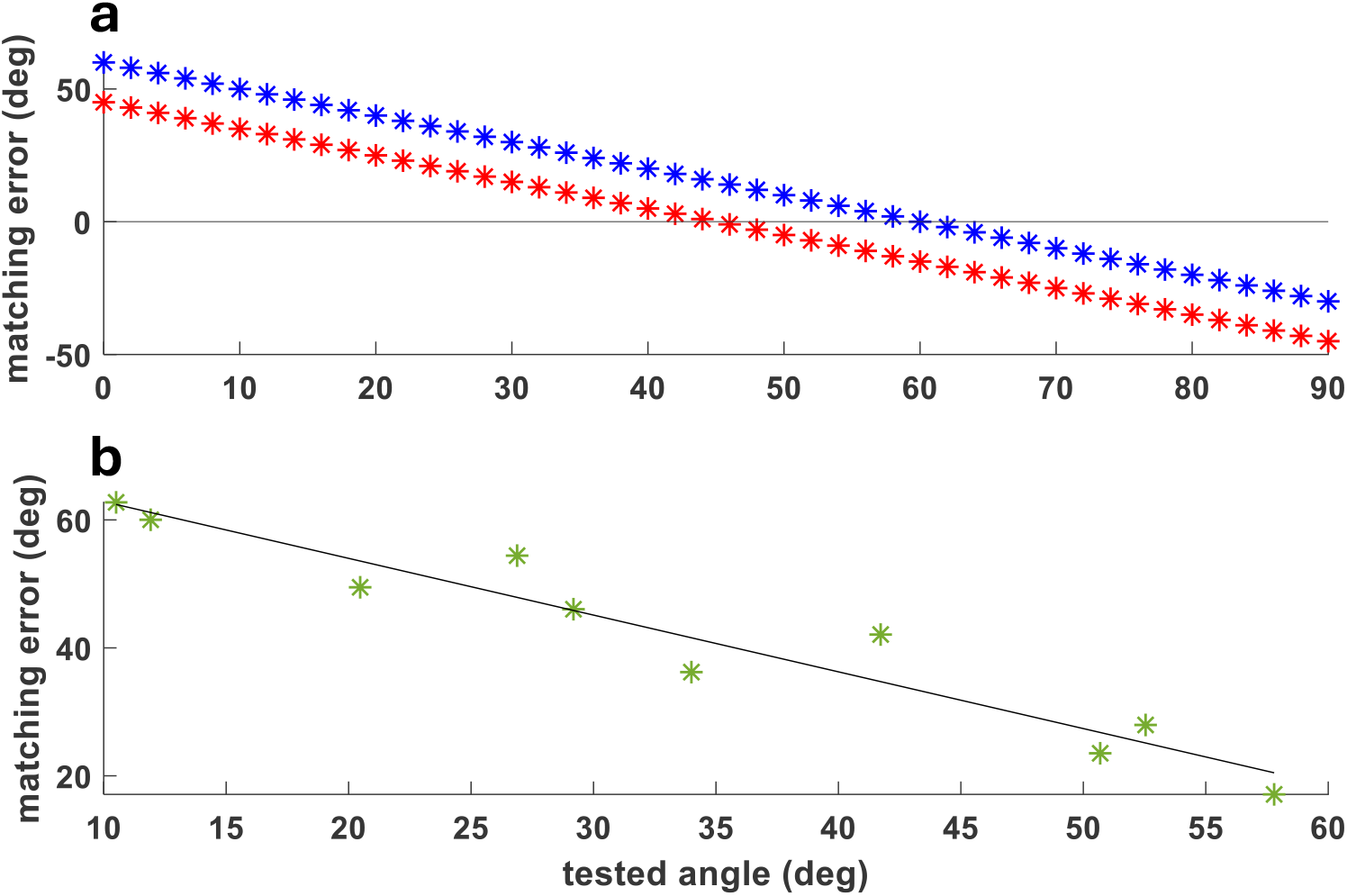
In panel **a**, hypothetical results from two SSPs tested across the 0–90° range in 2° intervals. One SSP operated under an expected prior of 45° (red), representing the mean of all tested angles, while the other used a prior of 60° (blue). In both cases, the linear relationship between error and tested angle had a slope equal to −1, with the intercept corresponding to the respective prior. In panel **b**, the observed results from a participant who reported no awareness of his arm’s position with eyes closed. The slope of the linear relationship was approximately −0.9, with an intercept near 70°.

A reasonable question arises: how could this person predict the structure of the test unless it is explicitly explained beforehand? To address this, we next asked, “What would this person do in the absence of such knowledge?” We reasoned that, lacking direct sensory input, the individual might rely on prior beliefs or assumptions about the test’s structure to guide their responses.

If the tested angles follow a prior distribution {*p*_*a*_}_*a*_ over the possible angle set *A*, the expected squared error is given by:

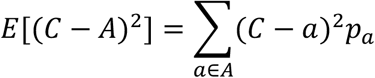

This expression represents a weighted version of the previous sum, where the uniform average is replaced by a weighted average using the prior probabilities {*p*_*a*_}_*a*_. To minimize this quadratic function with respect to *C*, we differentiate and set the derivative equal to zero:

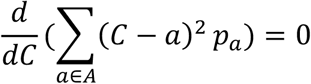

Solving gives:

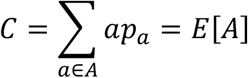

Thus, the optimal guess for *C* is the expected value of the set *A* under the prior distribution {*p*_*a*_}_*a*_. In simple terms, the optimal strategy would be to report the expected value of the prior (i.e., its mean).

In this case, the linear relationship between errors and tested angles is preserved, but the constant term becomes 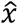, where 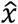 is the estimate of the prior distribution (i.e., *E*[*A*]). As a result, Accuracy becomes biased toward the prior, and both Precision and MAE (as well as RMS) continue to depend on the range of tested angles, as well as on the prior distribution. This effect is illustrated in Figure 1a (blue series), which shows how the error pattern shifts in response to prior assumptions.

This hypothetical person (i.e., SSP) illustrates that common performance measures in the position-matching task cannot be reliably interpreted as indicators of proprioceptive acuity since individuals with identical levels of sensory deficit (“senseless”) may receive different scores depending on their prior beliefs and the structural design of the test.

Given these predictions from the SSP model, using a data-driven approach to establish a threshold between healthy and pathological performance would require a comprehensive database. Such a database would need to include performance data for every joint, collected under a standardized test structure (using the same set of angles across all tests) with participants explicitly informed of the test parameters in advance. To the best of our knowledge, no such database currently exists, and constructing one would pose significant challenges. Importantly, many patients experience a limited range of motion due to factors such as pain, spasticity, or joint restrictions, which can prevent them from completing tests that rely on a fixed set of angles.

To address the challenges outlined above, and drawing from the strategic optimal responses (“guesses”) of the SSP, we propose the Guess Index (GI) as a novel outcome measure for proprioceptive acuity.

Given a set of tested angles {*x*_*i*_}_*i*_, the optimal performance of the SSP yields a predictable pattern of errors, as previously described. The predicted performance can be represented by the sum of the squared errors, 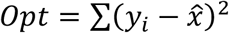as a single scalar. For this person, the observed sum of squares *Obs* = ∑(*y*_*i*_ − *x*_*i*_)^2^ is expected to be equal to the sum of squares of the optimal guess, 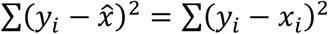. In cases other than complete loss of the sense, we expect *Obs*<*Opt*, so the ratio

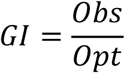

indicates whether the observed performance is equal to an optimal “senseless” guess (=1) or better (<1). The closer GI is to 0, the more sensory input available, with GI=0 indicating perfect sensory acuity. Naturally, GI can be >1, indicating performance worse than an optimal “senseless” guess. This new outcome ensures that the SSP would always get a score of 1, making it insensitive to the test’s structure or prior belief, as these are accounted for within the predicted optimal guess.

### Empirical Evidence

To demonstrate the utility of this approach, we collected data from healthy controls and individuals with suspected proprioceptive deficits using a clinically-oriented position-matching task. We then compared group performance across the various outcome measures.

#### Participants

Although not explicitly stated earlier, in the position-matching task, the SSP is assumed to have lost sensory input in one limb while retaining perfect sensory acuity in the contralateral limb, an essential condition for performing optimally in the task. Stroke is a common medical condition that frequently affects the sensory system [16–18], typically resulting in unilateral damage. This makes persons with stroke (PwS) ideal candidates for this investigation.

Participants who had experienced their first-ever stroke and had been independently performing activities of daily living prior to the event were recruited from an inpatient rehabilitation center (Adi-Negev Nahalat-Eran Rehabilitation Hospital). Exclusion criteria included the presence of other neurological conditions, mental instability, a history of substance abuse, or medical instability. Prospective participants were also excluded if they had orthopedic issues involving the arms, neck or torso.

Healthy controls were recruited from among the hospital staff, volunteers, and visitors. All control participants reported being free of any condition that could affect sensory acuity, including peripheral or central neurological disorders and orthopedic issues involving the arms, neck or torso. We recruited both younger (YC, <40 years) and older (OC, >50 years) control participants.

The study was approved by the Local Ethical Committee (ADINEGEV-2023_106), and all participants provided a written informed consent prior to enrollment.

#### Position-matching task

For clinical purposes, the test should be brief and easy to administer. In this study, we employed a simplified version of the position-matching task focused on the shoulder joint (encompassing both the glenohumeral and scapulothoracic components). Specifically, a physical therapist passively moved one shoulder of a blindfolded, seated participant to an arbitrary angle between approximately 10° and 90° of abduction. The participant was then instructed to replicate that angle using the contralateral shoulder and to verbally indicate when they believed the best match had been achieved. After each trial, the tested shoulder was returned to its anatomical resting position (approximately 0° of abduction). This procedure was repeated 10 times.

For PwS, testing was limited to the pain-free and available range of motion, including instances where restrictions stemmed from non-pain-related factors. In PwS, the paretic arm was designated as the index arm (i.e., the arm passively positioned by the examiner), whereas in control participants, the non-dominant arm served as the index.

#### Equipment and Processing

To quantify performance in the position matching task, two inertial measurement units (IMUs; Xsens DOT, Movella) were placed just above the elbows on the distal humerus of each arm, and a third unit was positioned on the back, between the shoulder blades. These IMUs recorded 3-axis gyroscopic, 3-axis accelerometric, and 3-axis magnetometric data at a sampling rate of 60 Hz.

The raw data were converted into earth-fixed angular measurements using the manufacturer’s proprietary algorithm, which has a declared sensitivity of 0.5 degrees. From these angular measurements, abduction angles were computed in two coordinate frames:

1. Elevation (absolute) angles – representing the arms’ orientation relative to the global, earth-fixed coordinate system.
2. Anatomical angles – representing the arms’ orientation relative to the angle of the back.

Under optimal testing conditions, where participants are seated upright with the main axis of the back aligned with gravity, no difference between these two coordinate frames is expected.

#### Additional tests

PwS were assessed using the upper-limb section of the Fugl-Meyer Assessment (FMA; [6]), a tool designed to measure impairment across multiple domains, including motor function, sensory ability, and coordination. Assessments were conducted by a licensed occupational therapist with expertise in administering the FMA.

#### Outcome measures

As outcome measures, we used the angular data to compute:

1. Accuracy= 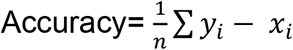
2. MAE= 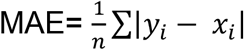
3. 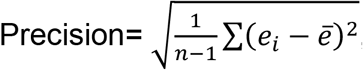, with 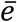 being Accuracy and *e*_*i*_ is the magnitude of the *i*th error. In simple terms, this is the consistency of the errors, as quantified by their standard deviation.
4. 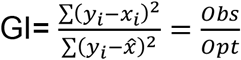, as explained above. As an estimate of 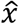, we used the mean of all the reported angles (*y*_*i*_).

These were computed for each coordinate frame (absolute and anatomical angles) separately.

#### Statistical analysis

To compare outcomes between groups and coordinate frames, we employed mixed-effects models, with each outcome variable treated as the dependent variable. Fixed effects included ‘Group’, ‘Coordinate Frame’, and the ‘Group-by-Coordinate Frame’ interaction, while ‘Subject’ was modeled as a random effect. When post-hoc pairwise comparisons were required, we used t-tests with sequential Bonferroni correction to account for multiple comparisons. Values for MAE and Precision were log-transformed to achieve a normal distribution of the models’ residuals.

For participants’ classification (Control vs. Patient), younger and older controls were combined into a single group. Logistic regression was then applied, with ‘Group’ as the predicted term and each performance metric as a predictor.

All analyses were conducted using the Statistical Package for Social Sciences (SPSS, version 29). The significance level was set a priori at α < 0.05.

## Results

A total of 9 younger controls (YC), 12 older controls (OC), and 15 PwS were recruited. Data from 2 OC participants were excluded due to technical issues. Participants’ demographics are summarized in Table 1.

**Table 1.** Participants’ demographics (median [IQR])

|  | Younger (n=9) | Older (n=10) | Stroke (n=15) |
| --- | --- | --- | --- |
| Age years | 28 [26-32] | 67 [63-68] | 68 [60-75] |
| Height cm | 170 [163-178] | 164 [151-173] | 167 [164-170] |
| Weight kg | 74 [60-82] | 67 [60-80] | 75 [65-83] |
| Sex F/M | 5/4 | 7/3 | 3/12 |
| FMU motor |  |  | 30 [21-53] |
| FMU sensory |  |  | 10 [9-12] |

*FMU is the Fugl-Meyer Assessment score (motor and sensory sections). 'F' and 'M' stand for Females and Males, and 'cm' and 'kg' stand for centimeters and kilograms.*

Descriptive statistics of task performance are presented in Table 2. A graphical representation of individual scores—based on elevation angles—is shown in Figure 2. Overall, test durations were brief (mostly under 2 minutes), and all participants successfully understood the instructions provided.

**Table 2.** Summary statistics of participants’ performance in the test.

|  | Younger (n=9) | Older (n=10) | Stroke (n=15) |
| --- | --- | --- | --- |
| Anatomical Angles (median [IQR]) |  |  |  |
| Accuracy | -34 [-42-(-16)] | -39 [-53-44] | 13 [-27-57] |
| MAE | 41 [34-66] | 50 [43-66] | 35 [22-57] |
| Precision | 6.5 [4.0-8.3] | 7.6 [6.3-9.2] | 9.7 [5.6-18.5] |
| GI | 0.92 [0.88-0.97] | 0.91 [0.90-0.94] | 0.97 [0.77-1.05] |
| Elevation Angles (median [IQR]) |  |  |  |
| Accuracy | 11 [1-16] | 8 [4-11] | 16 [-3-28] |
| MAE | 16 [11-20] | 9 [6-11] | 16 [11-28] |
| Precision | 4.7 [3.6-5.9] | 6.4 [5.7-8.2] | 7.9 [5.5-13.5] |
| GI | 0.58 [0.45-0.68] | 0.30 [0.23-0.51] | 0.90 [0.57-1.03] |

**Figure 2.**
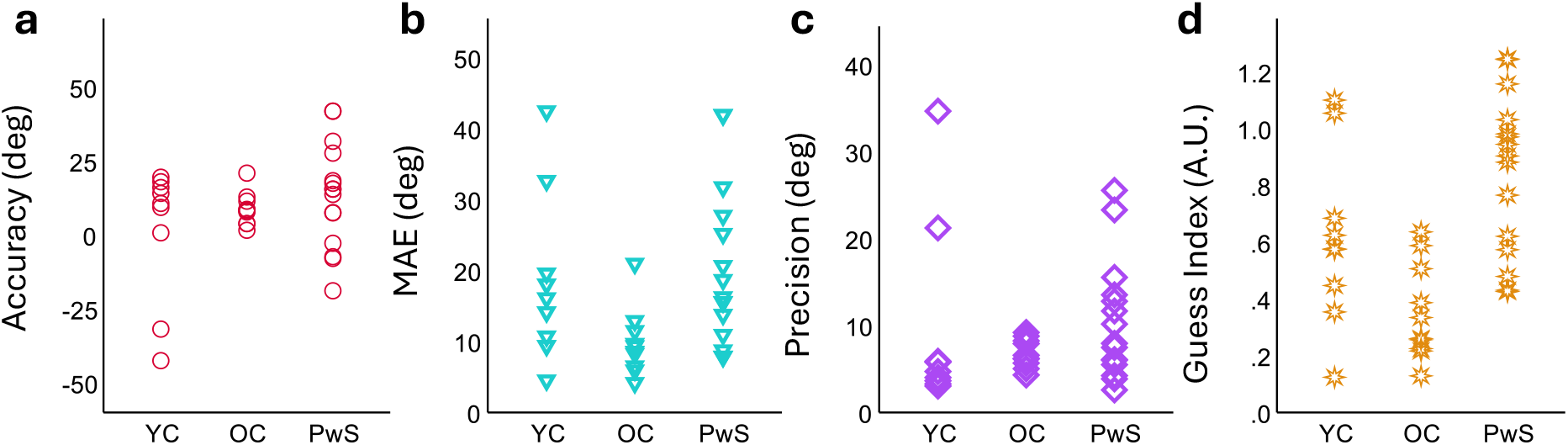
the score of each participant (in the elevation angles coordinate frame) captured by Accuracy (**a**), MAE (**b**), Precision (**c**) and GI (**d**). Two YC clearly performed at the level of the worst-performing PwS, in all measures.

A significant (p=<0.006) main effect for the coordinate frame was found in all models, excluding the Accuracy model. Results indicate that values in the elevation angles coordinate frame were lower than those observed in the anatomical angles coordinate frame.

A significant main effect for Group (p=0.012) was observed only in the GI model. Post-hoc comparisons revealed that values observed for OC were significantly (p=0.01) lower than those of PwS, but no other differences were observed.

A significant (p<0.001) interaction term was observed for the MAE and GI models. For MAE, exploring this term revealed a significant difference between OC and PwS only in the elevation angles coordinate frame. For the GI model, exploring this term revealed a significant difference between all groups (p<0.04), where the values for OC were the lowest, followed by YC, and the highest values were observed in PwS. Like the MAE model, these differences were only seen for the elevation angles coordinate frame.

Figure 2 reveals that two YC performed poorly, with scores comparable to the worst-performing PwS, and in some cases, even worse. When these two outliers are excluded, the performance of OC appears similar to that of the remaining YC group. Conversely, several PwS achieved scores within the range of healthy controls, suggesting that their stroke may not have significantly impacted proprioception.

Next, we tested whether any of the outcome measures could reliably classify participants as PwS. Since the elevation angle coordinate frame consistently yielded lower values and was the only coordinate frame showing significant group differences, we limited this analysis to elevation angles. For classification purposes, OC and YC were combined into a single control group. This binary grouping (Controls vs. PwS) served as the dependent variable in a logistic regression, with each performance measure entered as a predictor.

This analysis identified the GI as the only measure capable of classifying participants as PwS above chance level, achieving 80% accuracy (p = 0.004; see Figure 3).

**Figure 3.**
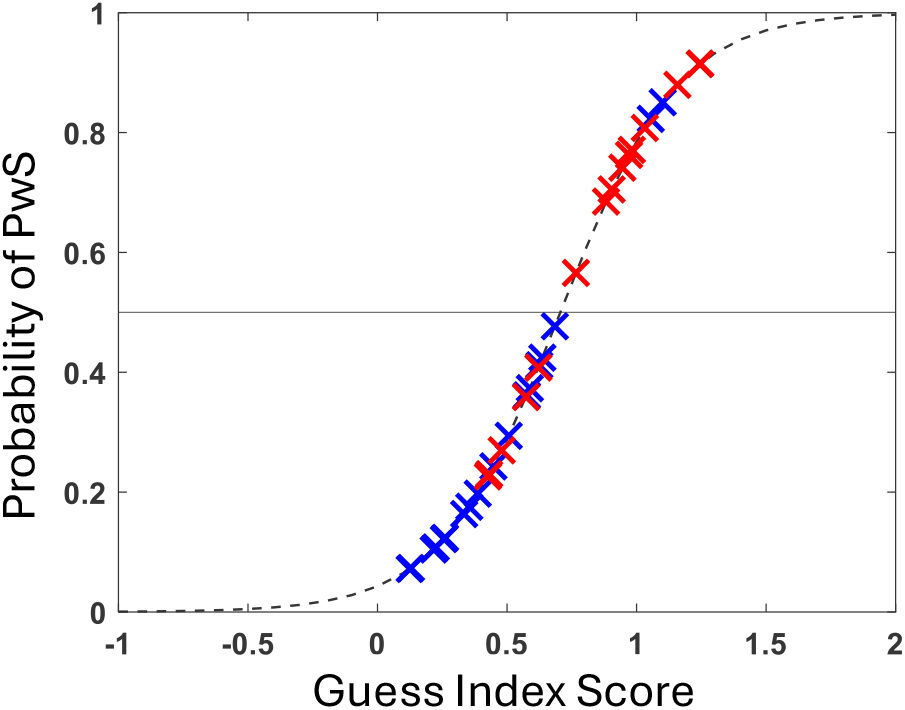
results of the GI logistic regression. Red X are PwS and Blue X are healthy controls. The horizontal line (probability=0.5) is the resultant threshold, indicating that participants with GI scores above ~0.7 were classified as PwS.

Next, we examined individual cases. Two PwS scored 4/12 on the sensory section of the FMA. One of these participants explicitly stated that, with eyes closed, he could not determine the position of his arm in space. The GI scores for these individuals were remarkably close to the optimal “senseless” guess level (0.98 and 0.97). Figure 1b illustrates the performance of one of these participants.

Six other PwS also performed at or above the optimal “senseless” guess level, despite having sensory FMA scores of 10 or 12—indicating minimal or no sensory deficit. A closer examination revealed that two of these participants scored 4 out of 36 on the upper extremity motor subsection of the FMA. Two others scored 29 out of 36, and the remaining two had full scores (36). Both participants who scored 29 reported shoulder pain during movement (Section J of the FMA), and one of them also scored zero on the coordination subsection. One of the participants with a full motor score reported a burning sensation along the affected arm, unrelated to movement.

## Discussion

The main findings of this investigation highlight that common performance measures in the position-matching task are sensitive to factors that are unrelated to sensory input. Using a combination of theoretical and empirical approaches, we demonstrated that these measures are influenced by the perceived objective and structural properties of the task, as well as by the prior beliefs and strategic responses of the tested individual. This sensitivity undermines the validity of these performance measures as indicators of sensory acuity. Building upon our theoretical framework, we developed a new outcome measure that is robust to these confounding factors and provided empirical evidence supporting its superiority over commonly used metrics. Below, we discuss the main findings, limitations, and implications of this work, and provide recommendations for interpreting the GI score.

First, we proposed that a threshold for proprioceptive deficits could be determined using a computational approach. In a simple example, we showed that a joint angle error of 2° would result in an end-effector error of approximately 1 cm. These values were not chosen arbitrarily.

The finger-to-nose test [19] typically requires an end-effector accuracy of approximately ±1 cm. Given the complexity of the upper limb—from shoulder to fingertip—comprising multiple joints and segments, it is unlikely that joint angle errors would naturally exceed 2°. Mean values far exceeding this are commonly reported [10,11,20–22], and it is presumed that individual errors, sometimes depicted in figures (e.g.,[23]), are even larger. Moreover, reported errors in end-effector positioning are often smaller than what would be expected based on the magnitude of individual joint errors [10,22]. This discrepancy likely indicates that consciousness has limited access to proprioceptive signals (see [24]), and/or that test responses are influenced by other factors related to the subjective nature of the task.

Alternatively, although the objective of the position-matching task might appear straightforward, our findings suggest that participants were more likely to perceive the task as matching the absolute angles of the limb segments rather than the anatomical angles of the shoulder, as reflected by lower matching errors within that coordinate frame. This result aligns with previous reports [13–15] and with the known contribution of gravity to the sense of body position, as reflected by the effect of microgravity [25]. The ambiguity regarding the coordinate frame likely contributes to the large errors reported in these prior studies. Nevertheless, we found that matching errors substantially exceeded our conservative estimate of 2° in both coordinate frames, suggesting that the CNS may not rely exclusively on either. In fact, there is an infinite number of possible coordinate frames that the CNS could adopt.

Regardless of the underlying cause, the magnitude of errors observed here, and those reported by others, imply that calculating a threshold based on a biomechanical model is not a feasible approach for detecting proprioceptive deficits in the position-matching task.

Second, an alternative approach to defining healthy sensory acuity thresholds is to empirically determine the performance limits of a healthy sensory system. While some researchers have adopted this strategy [21], inconsistencies in performance measures and methodologies across studies have impeded progress [26]. Moreover, our theoretical construct—the sensible senseless person—illustrates the inherent limitations of existing measures. Specifically, we showed that an individual with a fixed level of proprioceptive input can receive different scores depending on task structure, prior beliefs, and strategic responses (the latter is likely unconscious). This finding calls into question the validity of these measures. Importantly, such biases are not unique to individuals with deficits; due to noise in sensory input, prior beliefs are expected to influence position perception in all individuals [27]. For example, under any level of sensory uncertainty, if the tested angle exceeds the expected prior, the reported angle will be biased toward that prior, as negative errors are more probable [28].

Previous studies [22,23] have reported a linear relationship between error magnitude and target angles—consistent with our model’s predictions. Gritsenko et al. [23] attributed this trend to an optimal probabilistic strategy, while Fuentes and Bastian [22] interpreted it as evidence for signal-dependent noise, suggesting that conscious proprioception follows Weber’s Law. Regardless of the interpretation, this linear relationship indicates that test structure influences MAE, RMS, and Precision scores independently of sensory acuity, undermining their validity as measures of proprioceptive acuity.

Third, based on our theoretical framework, we developed the GI—a performance index designed to ensure that the SSP always receives a score equivalent to an optimal “senseless” guess, regardless of task structure or prior beliefs. The GI enhances interpretability by establishing a clear baseline for optimal guess performance (GI = 1), a feature absent in conventional measures.

Empirically, the GI demonstrated superior sensitivity relative to existing measures. Unlike traditional measures, the GI captured both group-level differences and individual-level variability. Specifically, it distinguished PwS from controls with an overall classification accuracy of 80% and uniquely identified individuals whose performance approximated optimal guessing—consistent with severe proprioceptive deficits documented in clinical reports and Fugl–Meyer scores. However, not all PwS who scored near 1 on the GI performed poorly on the sensory subsection of the FMA. Two such individuals had extremely low motor scores, and two others reported significant pain in the affected limb. Given that severe proprioceptive loss impairs motor control [1,2], it is reasonable to infer that these motor impairments may partially stem from sensory deficits undetected by the qualitative FMA. Pain may also influence proprioceptive perception (although this remains debated [29]), potentially explaining the high GI scores in individuals with substantial shoulder pain.

Interestingly, two younger controls also exhibited poor GI scores, suggesting that, like other measures, the GI is not immune to all cognitive influences. At the group level, these outliers caused the YC group to perform worse than the OC group— contrary to the typical effect of aging on sensory acuity [30] (although this effect might depend on the measure used [31]). Excluding these outliers (see Figure 2), the performance of the YC group was comparable to or slightly better than that of the OC group. At the individual level, the logistic regression classified these two participants as PwS, although sensory deficits were unlikely for these participants.

Accordingly, we propose that a GI score below 0.7 should be interpreted as an indication of adequate sensory input during testing, whereas scores above this threshold should be interpreted cautiously, as they may reflect sensory deficits and/or cognitive influence.

We do not claim that the GI is universally applicable in all contexts. For example, muscle conditioning [32] introduces systematic bias uniquely captured by Accuracy (and missed by Precision). Nevertheless, the GI’s strong theoretical foundation makes it superior to common measures for detecting proprioceptive deficits. Still, proprioception encompasses multiple dimensions beyond limb position; future work should aim to develop complementary measures that assess other aspects of this complex sense.

## Limitations

First, the subjective nature of the proposed task inherently exposes it to cognitive influences, regardless of the outcome measure. This was clearly evident in two healthy young controls. Although psychophysical paradigms, such as forced-choice designs [33], could mitigate such biases, they require specialized equipment and prolonged testing durations, limiting their clinical feasibility. Ultimately, an objective proprioceptive test is needed—an avenue our group is currently exploring.

Second, the relatively small sample size and the focus on a single anatomical structure and on PwS limit the generalizability of our empirical results, including the GI’s proposed threshold. We therefore encourage the research community to adopt this measure and to share original datasets, allowing the establishment of robust thresholds across joints and populations.

## Conclusion

Building on a simple version of the position-matching task, we developed a meaningful and theoretically grounded metric for unbiased quantification of proprioceptive acuity. Although we used the contralateral version of the test, the GI score can readily be applied to ipsilateral (position recall) or other task variants. The examination is brief, compatible with modern smartphones, and yields a simple, interpretable score derived from tested and reported angles. Our theoretical framework offers a strong foundation for constructing a standardized database of normative values, which may ultimately enhance the diagnosis and rehabilitation of sensory deficits.

## Data Availability

All data produced in the present study are available upon reasonable request to the authors

